# Decoding the impact of fibroblast heterogeneity on prognosis and drug resistance in high-grade serous ovarian cancer through tumor evolution analysis

**DOI:** 10.1101/2024.06.11.24308797

**Authors:** Tingjie Wang, Lingxi Tian, Bing Wei, Jun Li, Cuiyun Zhang, Ruitao Long, Xiaofei Zhu, Yougai Zhang, Bo Wang, Jun Yang, Yongjun Guo

## Abstract

**Background:** Tumor heterogeneity is associated with poor prognosis and drug resistance, leading to therapeutic failure. Here, we aim to utilize tumor evolution analysis to decode the intra- and inter-tumoral heterogeneity of high-grade serous ovarian cancer (HGSOC), unraveling the correlation between tumor heterogeneity and prognosis as well as chemotherapy response through single-cell and spatial transcriptomic analysis.

**Methods:** We collected and curated 28 HGSOC patients single-cell transcriptomic data from five datasets. Then, we developed a novel text mining-based machine learning approach to deconstruct the evolutionary patterns of tumor cell functions. This allowed us to identify key tumor-related genes within different evolutionary branches, elucidate the microenvironmental cell compositions that various functional tumor cells depend on, and analyze the intra- and inter-heterogeneity of tumors and their microenvironments in relation to prognosis and chemotherapy response in HGSOC patients. We further validated our findings in two spatial and seven bulk transcriptomic datasets, totally 1,030 patients.

**Results:** By employing transcriptomic clusters as proxies for functional clonality, we identified a significant increase in tumor cell state heterogeneity, which was strongly correlated with patient prognosis and treatment response. Furthermore, increased intra- and inter-tumoral functional clonality was associated with the characteristics of cancer-associated fibroblast (CAF). We also found that the spatial proximity between CXCL12-positive CAF and tumor cells, mediated through the CXCL12/CXCR4 interaction, is highly positively correlated with poor prognosis and chemotherapy resistance in HGSOC. Finally, we constructed a panel of 24 genes through statistical modeling, that are highly correlated with CXCL12-positive fibroblasts and can predict both prognosis and chemotherapy response in HGSOC.

**Conclusions:** Our study offers insights into the collective behavior of tumor cell communities in HGSOC, as well as potential drivers of tumor evolution in response to therapy. Functional analyses and experiments revealed a strong association between CXCL12-positive fibroblasts and tumor progression as well as treatment outcomes. Our findings provide an important theoretical basis for clinical HGSOC treatment.

## Introduction

Ovarian cancer (OC) ranks first in mortality among gynecologic malignancies [1]. High-grade serous ovarian cancer (HGSOC) is the most common and deadly subtype of OC [2]. Over 75% of HGSOC patients are diagnosed at an advanced stage with extensive malignant ascites and omental metastasis [3]. Complete staging surgery followed by platinum-based chemotherapy is the standard treatment for HGSOC [4]. However, over 25% of patients develop chemotherapy resistance within 6 months of initial treatment, and 70% experience recurrence within 2-3 years, ultimately succumbing to acquired drug resistance [5]. Therefore, platinum drug resistance is the primary cause of poor prognosis, recurrence, and death in HGSOC patients. Identifying clinical indicators that are closely associated with platinum chemotherapy resistance and HGSOC treatment prognosis is of great importance.

Tumor heterogeneity is a crucial factor influencing patient prognosis and treatment outcomes, which consists of inter-tumor (tumor by tumor) and intra-tumor (within a tumor) heterogeneity [6]. Intertumor heterogeneity mainly from different patients which induced by diverse genetic mutations, epigenetic modifications and transcriptional alterations. While, intra-tumoral heterogeneity refers to the presence of different cell types with varying functional characteristics within a tumor, resulting from interactions between tumor cells and distinct tumor microenvironment (TME) [6].

Previous research has primarily focused on intertumoral heterogeneity, analyzing genomic characteristics to obtain molecular subtypes of tumors, identify different patient subgroups, and select targeted therapies. For instance, four molecular subtypes have been identified in HGSOC: mesenchymal, immunoreactive, differentiated, and proliferative [7]. Among these, the immunoreactive subtype demonstrates a better prognosis and treatment response [7]. Identifying different mutational characteristics is crucial for treatment selection in patients. The BRCA1/2, TP53, and genes in the PI3K/AKT/mTOR pathway, which are frequently altered in ovarian cancer, serve as prime targets for clinical diagnosis and therapy [8]. However, intra-tumoral heterogeneity cause drug resistance, leading to therapeutic failure [6]. Thus, we need to integrate molecular features of both inter-tumor and intra-tumor heterogeneity with functional heterogeneity to improve patient subclassification and response to therapy.

Tumor heterogeneity is thought to adhere to the fundamental principles of Darwinian evolution, where individual cells harboring heritable mutations that enhance adaptability gain a survival advantage [9]. Natural selection drives clonal expansion, leading to the emergence of subclones with varying proliferative, migratory, and invasive capabilities [10]. The evolution of adaptive clones occurs within the dynamic tissue environment known as the tumor microenvironment (TME), forming a complex local tumor ecosystem. Changes within the TME further influence genetic diversification and phenotypic outcomes, driving the compositional tumor heterogeneity [11–13]. Therefore, identifying the distinct tumor cell clone subtypes within HGSOC and the microenvironment cell types they interact with, exploring the interactions between the microenvironment and tumor cells, and elucidating the patterns of tumor evolution are crucial for the treatment and prognosis prediction of HGSOC.

The TME composed of stromal cells, immune cells, extracellular matrix, and a variety of soluble factors, can influence tumor progression and response to therapy [13]. The utilization of single-cell and spatial transcriptomics technologies has significantly advanced the investigation of heterogeneity in HGSOC [7, 14]. For example, the presence of tumor-associated macrophages (TAMs) has been associated with poor prognosis in HGSOC, as these cells can promote angiogenesis, immunosuppression, and chemoresistance [15]. TGF-β-driven cancer-associated fibroblasts, mesothelial cells and lymphatic endothelial cells predicted poor outcome, while plasma cells correlated with more favorable outcome of HGSOC [7].

It becomes increasingly evident that understanding the evolutionary trajectories of tumor and TME cells is paramount to developing more effective and personalized treatment strategies [16]. It should explore the multifaceted intra-tumor heterogeneity and the role of the tumor microenvironment in shaping the evolutionary dynamics of ovarian cancer [17]. Understanding the biodiversity and clonality could provide conceptual knowledge about evolving tumor cell community with the ultimate goal of improving patient outcomes.

## Materials and methods

### Preprocessing of Single-Cell RNAseq Data

Raw gene expression matrices were processed by Seurat R package (version 3.2.3) [18]. Cells of low quality were filtered out based on two criteria: 1) Cells with fewer than 1000 unique molecular identifiers (UMIs) or with less than 100 genes detected; 2) Cells with more than 30% of their UMIs originating from mitochondrial genes. For each sample, gene expression matrices were normalized using the LogNormalize method via the NormalizeData function, and highly variable genes were identified using the scran package [19]. Dimensionality reduction was performed with Principal Component Analysis (PCA), where the number of principal components was selected based on the JackStraw function. Cell clusters were identified using the FindClusters function and visualized through Uniform Manifold Approximation and Projection (UMAP). Differential genes between clusters were calculated by the Wilcoxon rank-sum test, with a family-wise error rate set at 5%.

### Identification of Tumor Cells

Copy number variations (CNVs) were estimated in each individual cell by analyzing the averaged expression profiles across chromosomal intervals. The initial estimation of CNVs for each chromosomal region was conducted using the infercnv R package (version 1.0.4)(inferCNV of the Trinity CTAT Project. https://github.com/broadinstitute/inferCNV). In epithelial cells, CNVs were determined by comparing the expression levels derived from the single-cell RNA sequencing (scRNA-seq) dataset, with a cutoff value of 1 and a noise filter set at 0.2. For each sample, gene expression data were re-standardized, and the values were constrained within the range of -1 to 1. The CNV for each cell was then calculated as the sum of the squares of the CNV values across the respective regions.

### Calculation of Epithelial Cells Scores

We employed the expression levels of epithelial scores based on the expression of five well-established markers: *KRT19*, *KRT7*, *KRT18*, *KRT8* and *EPCAM*, as reported in previous studies [20].

## Tumor Evolution Analysis

### Selection of Representative Tumor Cells

Input the clustering information from Seurat, and simultaneously identify the tumor cell subgroups using the previously mentioned CNV method.

Utilize the normalized matrix to calculate the Principal component (PC) scores for each cell. Subsequently, employ the Wilcoxon rank-sum test to obtain PCs that show significant differences between classes (FDR < 0.05). Finally, select the top 10 genes with the highest absolute coefficients of the differentially expressed PCs as the characteristic genes for the corresponding principal components.

Employ the characteristic genes obtained from the tumor subgroups, and then apply the TF-IDF (Term Frequency-Inverse Document Frequency) method. This technique is typically used in information retrieval and text mining to evaluate how important a word is to a document in a collection or corpus. In the context of gene expression analysis, TF-IDF can be used to identify the most relevant genes in the tumor subgroups based on their frequency and uniqueness across the dataset.

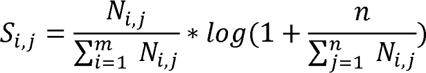

where N_i,j_ is the m by n normalized gene expression matrix defined above. Within the TF-IDF method, set the lower quartile of a gene’s expression across all cells as the threshold. If a gene’s expression is below this threshold, set its expression to 0. This approach ensures that only genes with a certain level of expression are considered significant, effectively filtering out genes that are expressed at very low levels across the cells. After applying the TF-IDF method to obtain the gene weight matrix S_i,j_, calculate the 95th percentile (A) of each gene’s weight across all cells and set the lower threshold for gene weights at 0.25 * A. Any gene weight below this threshold in S_i,j_ is set to 0, resulting in a corrected S_i,j_ matrix. Subsequently, for each cell group, sum the weights of the characteristic genes to derive a total weight and set the cutoff value T to establish the cell weight threshold utilizing the formula:

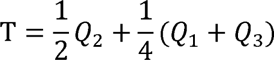

where Q1, Q2 and Q3 represent lower quartile, median and upper quartile value separately. Cells with score higher than T are identified representative tumor cells.

### Clustering and refinement

Utilizing the high-purity tumor cells obtained in the first step for each tumor subgroup, we employed the SC3 method for consensus clustering to obtain subclassification labels for each subgroup. Subsequently, we merged the subcategories of each subgroup to identify those with statistically significant differences. Specifically, we first extracted the top 1000 highly variable genes from the PCA of high-purity malignant cells. For each subgroup’s subclassification obtained through consensus clustering method SC3 [21], we trained a support vector machine (SVM) model using the first 10 principal components and validated the classification accuracy through ten-fold cross-validation. Following this, we randomly shuffled the labels to perform a permutation test (randomly shuffled 100 times), which allowed us to calculate the classification significance P-values between each pair of clusters. These P-values were then sorted in descending order, and an iterative merging process was conducted until the largest P-value was less than 0.05, at which point the iteration ceased. Through this method, we were able to identify the subclusters within each tumor subgroup that were of the highest purity and had statistically meaningful classifications.

### Hierarchical evolutionary analysis

We intersect the features derived from TF-IDF with those obtained through PCA and obtain tumor-associated feature genes. Utilizing these genes, above high-purity tumor cells and the purified subpopulations of each subgroup, we compute the mean expression values of the corresponding subgroup’s feature genes to generate a pseudo-bulk dataset. We then apply the removeBatchEffect function in the Limma package (version 3.58.1) to remove for batch effects across various datasets. Subsequently, we perform hierarchical evolutionary analysis between subgroups using the pvclust function from the pvclust package (version 2.2, nboot=1,000).

According to the aforementioned step, we are able to obtain highly purified subpopulations within tumors, facilitating the analysis of intratumoral heterogeneity. Additionally, this method allows us to establish hierarchical evolutionary relationships among samples to obtain BR1, BR2 and BR3, thereby shedding light on their intertumoral heterogeneity. Importantly, it also enables the identification of genes that are indicative of the tumor characteristics specific to the entire ovarian cancer population.

### Cell Type Enrichment Analysis

Each cell subtype encompassed a range of tumor stages, for which we computed enrichment scores (EScores) to quantify their prevalence at different stages. These EScores, reflecting the ratio of subtype cell numbers at specific stages to their overall distribution, highlighted when a subtype was predominantly enriched, with values greater than 1 signifying enrichment at that stage [20].

### WGCNA analysis in cell subclusters and acquisition of fibroblast-related Genes

The normalized expression matrix was utilized to construct a weighted gene co-expression network via the WGCNA R package (version 1.69). To mitigate the impact of noise and outliers, analysis was carried out on ‘pseudo cells’, which represent the average gene expression of ten randomly selected cells within each distinct cell type [22]. Network construction was achieved using the ‘blockwiseModules’ function, applying the default settings. For each identified module, a principal component analysis was performed using the module eigengenes. The correlation between module eigengenes and cell type metadata was calculated to evaluate the relevance of each module using Pearson’s correlation test. Subsequently, hub genes within significant modules were identified based on their modular connectivity, which refers to the absolute value of Pearson’s correlation between genes (module membership), and their relationship with clinical traits, defined as the absolute value of Pearson’s correlation between individual gene expression and cell type. Perform WGCNA analysis in the fibroblast subpopulations of datasets GSE154600 and GSE165897 [23], and take the intersection of characteristic genes of cell subpopulations significantly infiltrating in the BR3 branch as the final fibroblast-related characteristic genes, which will be used for the construction of the risk prognosis and drug resistance analysis model below.

### Similarity analysis among cell subpopulations

Utilize the acquired single-cell subpopulations and the union of characteristic genes obtained through WGCNA. Calculate the mean expression levels of these genes in each subpopulation and sample phenotype (the evolutionary branch to which the sample belongs). Use the R package ggcor (https://github.com/hannet91/ggcor) to compute the correlations between cell subpopulations and between cell subpopulations and phenotypes, and visualize these correlations.

### Clustering of bulk RNA data samples

Bulk RNA data were retrieved from the Cancer Genome Atlas (TCGA; https://www.cancer.gov/tcga) and GEO dataset, respectively. Subsequently, NMF clustering methods were performed on the normalized expression data using the NMF R package (version 0.23).

### Mapping Analysis of Single-Cell and Bulk RNA Subpopulations

To establish the correspondence between the inter-tumor clusters obtained from single-cell evolutionary analysis and the clusters identified through clustering methods in bulk, we first extract the characteristic gene sets from the single-cell BR1, BR2, and BR3 subpopulations. Subsequently, we employ a hypergeometric test to calculate the enrichment scores of the bulk RNAseq samples within these three gene sets. We then select the connections with the highest enrichment scores that are significantly enriched as the mapping relationship between single-cell and bulk RNAseq samples.

### Friends analysis

The Friends analysis approach assesses the functional correlation among various genes within a pathway, suggesting that the interaction of a gene with others in the same pathway enhances its likelihood of expression. This methodology is widely adopted to identify crucial genes. Utilizing the R package GOSemSim [24], we calculated the functional correlations among genes linked to the prognosis of high-grade serous ovarian carcinoma (HGSOC) and drug resistance.

### Spatial Transcriptome Data Analysis

Raw gene-spot matrices were analyzed with the Seurat package (version 3.2.3) in R. Spatial transcriptome data were qualitatively controlled using parameters including total spots, media UMIs/spot, median genes/spot, median mitochondrial genes/spot. Spots used in the subsequent analysis were filtered for minimum detected gene count of 200 genes while genes expressed in fewer than 3 spots were removed (Supplementary Table 2). Normalization across spots was performed with the SCTransform function. Dimensionality reduction and clustering were performed with principal component analysis (PCA) at a resolution of 1 with the first 30 PCs. We conducted cluster analysis using FindClusters, and then utilized the standardized expression matrix to calculate the average expression levels of immune-related genes (*PTPRC*, *CD2*, *CD3D*, *CD3E*, *CD3G*, *CD5*, *CD7*, *CD79A*, *MS4A1*, *CD19*) [25]. The subpopulation with the highest expression levels was selected as the normal control. We employed the InferCNV method to identify subpopulations of tumor cells. Subsequently, using the RegionNeighbours function from the R package STutility (version 1.1.1, https://ludvigla.github.io/STUtility_web_site/), we determined the cells at the tumor edge. We then used the FindMarkers function to calculate the differentially expressed genes in the region adjacent to the tumor edge.

### Immune Infiltration Analysis

We assessed the immune score of various immune cells in HGSOC patients by employing xCell (R package, version 1.1) on RNA-seq datasets and microarray datasets [26]. The microarray datasets underwent quantile normalization, while the RNA-seq dataset was quantified in terms of FPKM.

### In Vitro Functional Assays

To investigate the role of the *CXCL12/CXCR4* signaling pathway in ovarian cancer, siRNA was used to knock down *CXCR4* in SKOV3 cells. The knockdown effects of different siRNA sequence were detected by qRT-PCR and Western blot respectively. The CCK8 assay was utilized to examine changes in cell viability after CXCL12 treatment and to determine the optimal cytokine concentration. Subsequently, cellular viability was monitored in the presence of the optimal concentration of CXCL12 to discern the variations induced by CXCR4 knockdown.

### Cell Culture

The human ovarian cancer cells SKOV3 were purchased from the ATCC cell bank. The cells were cultured in McCoy’s 5A medium supplemented with 10% FBS and 1% double antibiotics (100 U/mL penicillin, 100 g/L streptomycin), and incubated in 37 °C incubator with 5% CO_2_. Trypsin was used for digestion and passaging when the cell reached 80%-90% confluence.

### Cell Transfection

Cells were digested by trypsin and prepared into cell suspension. After cell counting, cells were plated at a density of 4×104 per well in a 24-well plate and incubated overnight in a cell culture incubator. siRNA transfection was performed according to the protocol of Lipofectamine 3000 (thermofisher L3000015), using 15 pmol siRNA and 1.5 μL Lipofectamine per well. After 48 hours incubation, cells were collected for qRT-PCR and Western blot. The sequences of siRNA for the knockdown and control groups are shown as following:

> si-CXCR4-1
>
> GGCAAUGGAUUGGUCAUCCUGGUCA
>
> UGACCAGGAUGACCAAUCCAUUGCC
>
> si-CXCR4-2
>
> UGGUUGGCCUUAUCCUGCCUGGUAU
>
> AUACCAGGCAGGAUAAGGCCAACCA;
>
> si-CXCR4-3
>
> UGUUUCCACUGAGUCUGAGUCUUCA
>
> UGAAGACUCAGACUCAGUGGAAACA;
>
> si-NC
>
> UUCUCCGAACGUGUCACGUTT
>
> ACGUGACACGUUCGGAGAATT

### qRT-PCR

qRT-PCR was used to detect the transcriptional level of the CXCR4 gene and knockdown effect of different siRNA sequences. TransZol Up was added to the cell samples for lysis, and the samples were processed according to protocol for RNA extraction. After air-drying at room temperature, RNA pellets were dissolved in 20 μL Rnase free H_2_O. Concentration and A260/280 of RNA samples were measured using a spectrophotometer. Reverse transcription was performed according to the First-Strand cDNA Synthesis SuperMix kit instructions, 1 μg total RNA was used as template. The reaction mixture was incubated at 50 °C for 5 minutes and 85 °C for 2 minutes. Then qPCR reaction system was prepared according to the PerfectStart® Green qPCR SuperMix instructions, and the real-time PCR program was set as following, holding stage step1 95.0 °C 30 sec; cycling stage : number of cycles 40, step1 95.0 °C 15 sec, step2 60.0 °C 30s; melt curve stage: 95.0 °C 15 sec, 60.0 °C 60 sec, 95.0 °C 15 sec. The primer sequences used were:

> CXCR4-h-F ACTACACCGAGGAAATGGGCT;
>
> CXCR4-h-R CCCACAATGCCAGTTAAGAAGA;
>
> GAPDH-h-F TGACAACTTTGGTATCGTGGAAGG;
>
> GAPDH-h-R AGGCAGGGATGATGTTCTGGAGAG.

### Western Blot

Western blot was used to detect the protein level of CXCR4. Cell samples were treated with RIPA lysis buffer (Sigma-Aldrich) containing PMSF and protease inhibitors, sonicated on ice, centrifuged at 4 °C, 12000 g for 10 minutes, and the supernatant was taken for protein quantification using the BCA method. After mixing with loading buffer and denaturing at 100 °C for 5 minutes, proteins were separated using 10% SDS-phage gel and then transferred to PVDF membrane. After blocking with 5% skim milk for 1 hour at room temperature, the membrane was incubated with the primary antibody, followed by HRP-conjugated secondary antibody, and imaged using ECL chemiluminescence.

### CCK8

The CCK8 assay was used to detect the effect of *CXCL2/CXCR4* on cell viability. cells were trypsinized to prepare a cell suspension When reached 80%-90% confluence. After cell counting, cells were plated at a density of 3000 cells per well in 96 well plate. After incubation in a 37 °C incubator for 24 hours, the medium was replaced with complete medium containing (0, 50, 100, 200, 300 ng/mL) CXCL12 and cultured for 48 hours. 10 μL of CCK8 reagent was added per well, and the absorbance at 450 nm was measured using a microplate reader after 1 hour incubation in 37 [. For further research, si CXCR4 and si NC transfections were performed at 24 hours respectively after plating. 6 h later, CXCL12 was added to a final concentration of 200 ng/mL, and the CCK8 assay was conducted using the same method.

## Results

### Construction of a Multi-Omics Atlas of HGSOC including Chemotherapy

#### Response Characteristics Using Single-Cell and Spatial Transcriptomics

To enhance the understanding of the complex interplay between intratumoral and intertumoral heterogeneity in high-grade serous ovarian cancer (HGSOC), and to develop a comprehensive tumor evolutionary model that delineates the distinct tumor and microenvironmental profiles across diverse patient cohorts, we have meticulously assembled a robust dataset (Figure 1A). This dataset comprises four single-cell datasets and two spatial transcriptomics datasets, totaling 50 samples. Notably, 28 of these samples (57%) were obtained from patients prior to chemotherapy treatment and exhibited measurable responses to therapy (Figure 1B). Complementing this, we have also curated two bulk RNA sequencing datasets that include chemotherapy treatment responses, encompassing 24 samples (Table S1). Our analytical approach began with the application of advanced text mining and machine learning algorithms to three of the single-cell datasets. This enabled the construction of a sophisticated hierarchical evolutionary model specific to HGSOC. This model was then rigorously validated and further scrutinized for insights into the intricate relationships between intratumoral microenvironmental heterogeneity and intertumoral population heterogeneity using the remaining two datasets. This step was crucial for establishing the model’s reliability and for uncovering potential links between cellular heterogeneity and therapeutic outcomes. Subsequently, we focused on the spatial transcriptomics datasets to dissect the spatial distribution characteristics of microenvironmental heterogeneity, providing a visual representation that enhances the interpretation of our findings. In the merged test dataset, which consisted of 14 samples, we processed and analyzed a total of 87,288 cells post-quality control (Figure 1C). These cells were classified into several key subtypes: B cells (5,700, 6.53%), CAF (cancer-associated fibroblasts, 17,664, 20.24%), ENDs (Endothelial cells, 1,174, 1.35%), EPI cells (Epithelial, 20,273, 23.22%), Mono cells (Monocyte, 15,217, 17.43%), and T_NK cells (T and NK cells, 27,259, 31.23%). Furthermore, we employed the CNV method and calculated the epithelial cell score to specifically identify 18,273 tumor cells (Figure 1C). Through this integrated approach, we aim to not only elucidate the evolutionary dynamics of HGSOC but also to provide a foundation for the development of personalized therapeutic strategies that account for the unique heterogeneity observed in each patient’s tumor.

**Figure 1.**
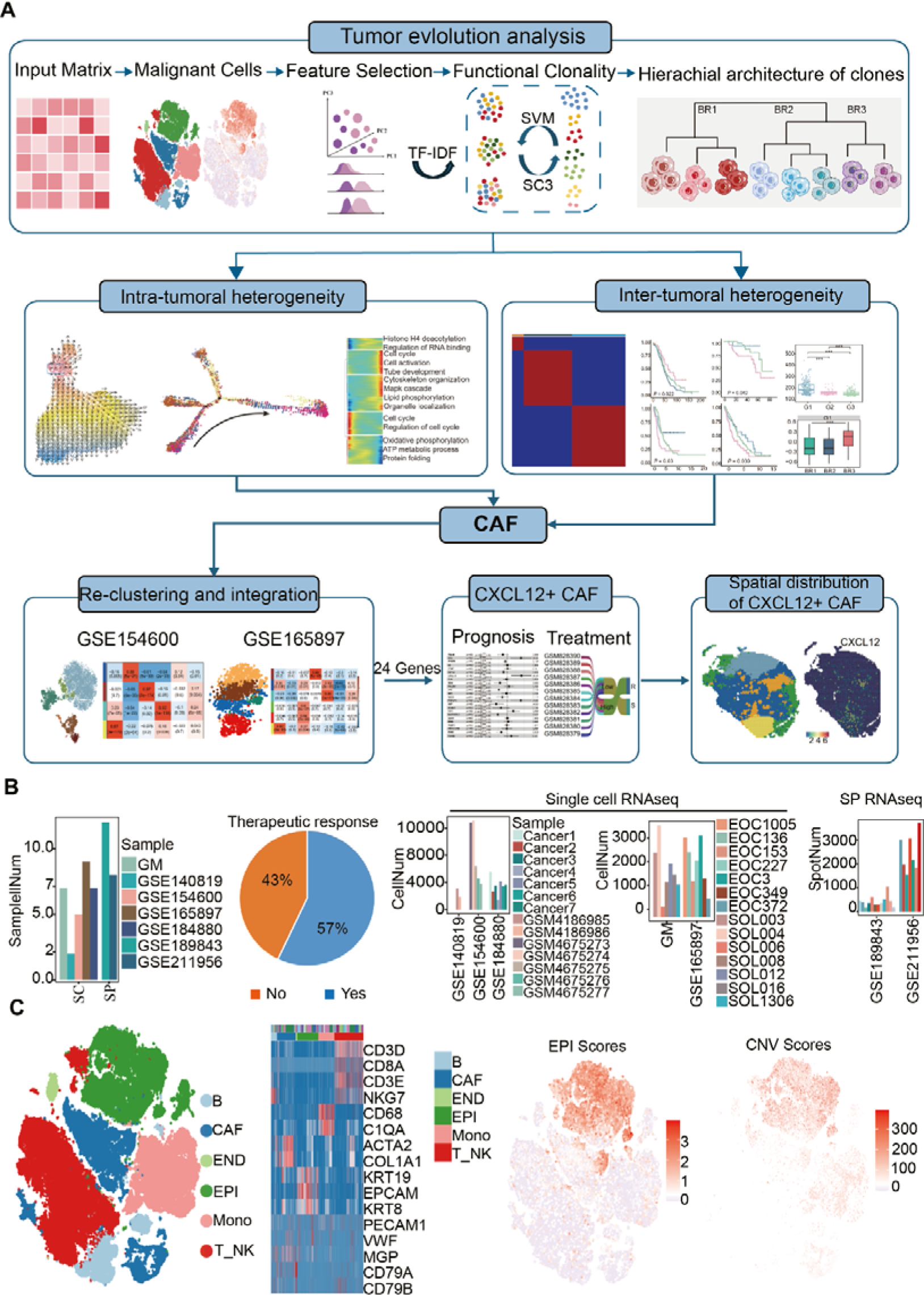
HGSOC transcriptome atlas. (A) Schematic depicting the study design. The number of samples in the tumor evolution analysis of HGSOC. Pie chart showing the proportion of clinical treatment in the tumor evolution analysis. The number of cells and spots scRNA-seq datasets. (C) The t-distributed stochastic neighbor embedding (t-SNE) plots showing the major cell types in HGSOC. Clusters are distinguished by colors. (D) Heatmap showing cell type marker genes expression level in the first single cell dataset. (E) Expression profile of epithelial and tumor scores in the first single cell dataset, the color from gray to red represents the expression level from low to high.

#### Study on Inter- and Intra-Tumoral Heterogeneity in High-Grade Serous Ovarian Cancer

Inter- and intra-tumoral heterogeneity significantly impacts patient prognosis and treatment response. We employed text mining and consensus clustering to dissect the clonal evolution within individual patients, followed by hierarchical clustering and bootstrap analysis to establish inter-tumoral clonal evolutionary relationships. Through this approach, we identified three evolutionary branches, designated as BR1, BR2, and BR3, across 69 clusters in 14 samples from test dataset 1 (Figure 2A, including samples in GSE140819, GSE154600 and GSE184880). Notably, individual tumor samples tended to cluster under a specific evolutionary branch. To validate the robustness of our method, we replicated the analysis in a subset of the data, confirming consistent evolutionary relationships among cell populations in dataset GSE184880 (Figure S1).

**Figure 2.**
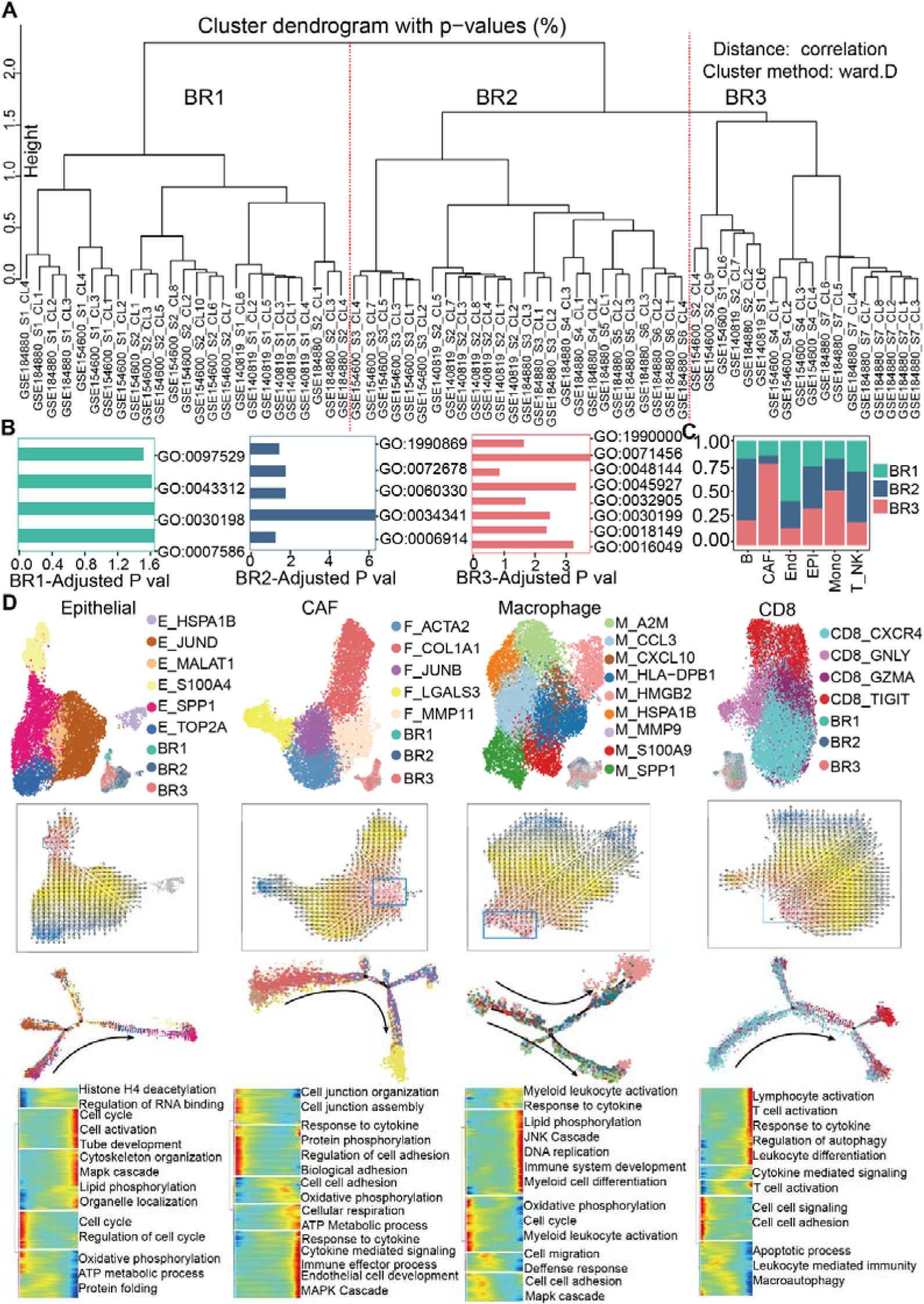
Tumor branching evolution reveals intratumor heterogeneity in HGSOC. (A) Tumor phylogenetic tree constructed by hierarchical clustering of all the clusters from 14 tumors, in which BR1, BR2 and BR3 were defined according to the hierarchical relationship. (B) Bar plot showing enrichment analysis using the tumor branch evolution features via clusterProfiler. (C) Bar plot showing the sample origins of three subtypes of branching evolution. (D) Distribution characteristics of intratumor cell types obtained through tumor evolutionary analysis. Profile and Uniform Manifold Approximation and Projection (UMAP) plots showing the sub cell type in the epithelial, CAF, macrophage and CD8 cells (top). Velocity and single-cell trajectory result (row 2 and row 3), Differentially expressed genes (rows) along the pseudo-time (columns) were clustered hierarchically into five groups in the scRNAseq dataset. Pathway enrichment scores were calculated using clusterProfiler.

Having established the clonal evolutionary relationships across different samples, we identified key genes within these branches and conducted functional analyses (Table S2). Key genes of BR1 were enriched in pathways related to myeloid leukocyte migration (GO:0097529), neutrophil degranulation (GO:0043312), and digestion (GO:0007586). BR2 genes were predominantly associated with cellular response to chemokine (GO:1990869), T cell migration (GO:0072678), regulation of response to type II interferon (GO:0060330), and autophagy (GO:0006914). BR3 genes were enriched in pathways such as the generation of amyloid fibrils (GO:1990000), cellular response to hypoxia (GO:0071456), positive regulation of growth (GO:0045927), and collagen fibril organization (GO:0030199) (Figure 2B). These findings suggest that BR3 is characterized by highly fibrotic, immune desert-like tumors, BR2 by T cell-infiltrated hot tumors, and BR1 by predominantly tumor cell-dominated cold tumors [27].

Further analysis revealed significant heterogeneity in the distribution of different microenvironmental cells across the branches, with CAF cells notably enriched in the BR3 branch (Figure 2C). To delve deeper into intra-tumoral heterogeneity, we dissected the heterogeneity of epithelial, fibroblast, monocyte, and CD8 cells. We identified six epithelial cell subpopulations (E_HSPA1B, E_JUND, E_MALAT1, E_S100A4, E_SPP1, E_TOP2A), with E_SPP1 and E_TOP2A predominantly found in the BR3 branch (Figure 2D). *SPP1* was shown to be highly expressed in multiple tumor types and interacts with fibroblasts or T cells via pathways such as SPP1-CD44, correlating with tumor malignancy. *TOP2A*, a cell cycle gene, indicates high proliferative states when overexpressed. Cell velocity and pseudo-time analysis revealed that epithelial cells evolve towards the BR3 subpopulation, with terminal pathways enriched in cell cycle, epithelial differentiation, and lipid metabolism (Figure 2D).

We identified five fibroblast subpopulations (F_ACTA2, F_COL1A1, F_JUNB, F_LGALS3 and F_MMP11), most of which were concentrated in the BR3 branch (Figure 2D). Cell velocity analysis showed enrichment of cell adhesion-related pathways at the onset and metabolic, chemokine, and MAPK pathways at the end (Figure 2D). Eight macrophage subpopulations were identified (M_A2M, M_CCL3, M_CXCL10, M_HLA-DPB1, M_HMGB2, M_HSPA1B, M_MMP9, M_SPP1), with M_CCL3, M_CXCL10, M_HLA-DPB1, and M_SPP1 enriched in the BR3 branch (Figure 2D). Temporal analysis revealed enrichment of lipid metabolism, granulocyte differentiation, and JNK-related pathways at the terminal stage (Figure 2D). Four CD8 cell subpopulations were identified, with CD8_GZMA and CD8_TIGIT enriched in BR1 and BR2, and BR3 dominated by CD8_CXCR4. Cell velocity analysis showed enrichment of T cell activation, lymphocyte differentiation, and autophagy regulation pathways at the terminal stage of cell differentiation (Figure 2D).

These results underscore significant inter- and intra-tumoral cellular heterogeneity in HGSOC, with epithelial cells in BR3 samples exhibiting high proliferation rates and infiltration by diverse fibroblast subtypes.

#### Tumor functional clonality is associated with patient prognosis and TME composition

In our previous analysis, we divided all single-cell samples into three branches and identified significant heterogeneity between these branches, along with 150 characteristic genes for each branch (Table S2). We further examined the prognostic clustering effect of these genes. Prognostic analysis across five datasets (TCGA, GSE14764, GSE26193 and GSE26712, Table S1) revealed a notably strong prognostic clustering effect in these datasets, with the poorest prognosis samples exhibiting high levels of fibroblast infiltration (Figure 3A, 3B, G1 in TCGA, G2 in GSE14764, GSE26193 and GSE26712). Correlation analysis also indicated a significant high correlation between fibroblast content and fibroblast-associated marker genes (*COL3A1*, *COL1A2*, *COL1A1*, *FN1* and *DCN*, Figure S2). Subsequently, we used the inter-tumoral heterogeneity branch genes obtained as a reference gene set to perform a hypergeometric test in different bulk samples, calculating the distribution characteristics of samples in the single-cell BR branches. The results showed that bulk samples with the poorest prognosis and high fibroblast infiltration were significantly enriched in BR3 scores (Figure 3C), further validating the reliability of using single-cell samples for studying inter-tumoral heterogeneity in tumors. Differential gene analysis in bulk samples also revealed enrichment of numerous pathways related to epithelial cell migration, cell adhesion, and chemokines in samples with poor prognosis (Figure 3D). These findings suggest that the content of fibroblasts in high-grade ovarian cancer samples may be highly correlated with tumor malignancy.

**Figure 3.**
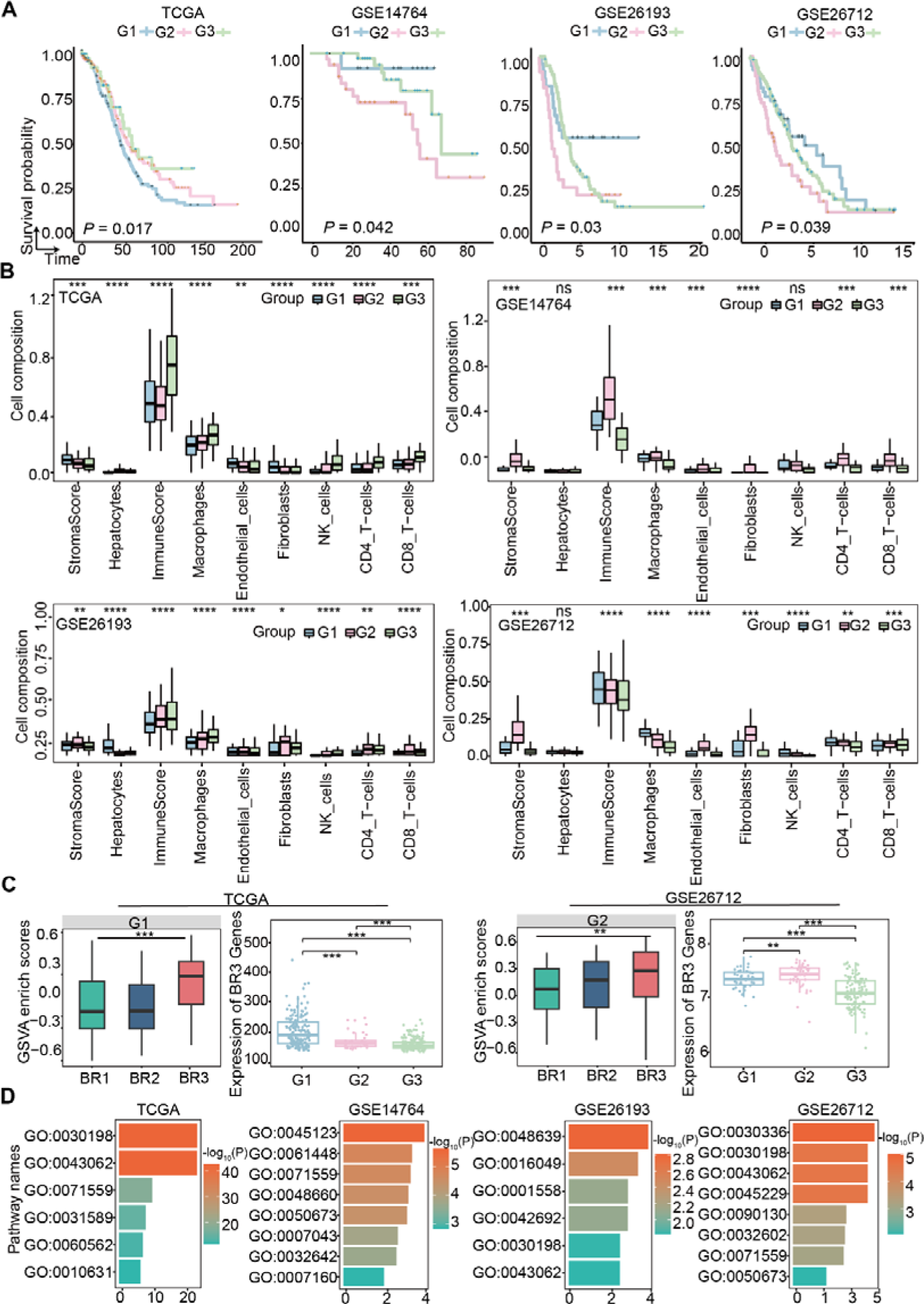
Tumor branching evolution reveals inter-tumor heterogeneity and proportion of fibroblasts promote the poor prognosis of HGSOC. (A) Overall survival curves showing the prognosis result of the three subtypes (G1, G2 and G3) obtained from NMF clustering using the 150 tumor evolution features in the TCGA and GEO cohorts. (B) Boxplots showing the immune cell infiltrates ratio in the three distinct malignant subtypes in the significantly enriched patients via xcell (ns, no significance, *P < 0.05, **P < 0.01, ***P < 0.001). Pairwise comparison was conducted by Wilcoxon rank-sum test in the RNA cohort. For the boxplot, the centerline represents the median and box limits represent upper and lower quartiles. (B) Boxplot showing the GSVA enrichment scores in the poorest prognosis using the branch features of tumor evolution analysis in scRNA datasets. Boxplots showing the mean expression level of BR3 genes in the three subtypes of bulk RNA datasets, ns, no significance, *P < 0.05, **P < 0.01, ***P < 0.001, Wilcoxon rank-sum test. (D) GO enrichment analysis of upregulated genes of the poorest prognosis group (G1 in TCGA, G2 in the other cohorts).

#### Identification of a subpopulation of CXCL12-positive fibroblasts associated with chemotherapy resistance and poor prognosis through intra- and intertumoral heterogeneity analysis of fibroblasts

Previous research has demonstrated a strong correlation between heterogeneity of TME composition, especially the infiltration of fibroblast, and prognosis. Given that chemotherapy is commonly employed in the early stages of ovarian cancer, we further explored the relationship between intra-tumor heterogeneity and chemotherapy resistance. To this end, we analyzed the differences in cellular composition of the tumor microenvironment in tumors with varying responses to chemotherapy. Utilizing the 150 characteristic genes obtained, we conducted a tumor evolution analysis on five samples within the single-cell dataset GSE154600, revealing that the chemotherapy-sensitive samples GSM4675276 and GSM4675276 belonged to the BR1 subgroup, while the chemotherapy-resistant samples GSM4675273 and GSM4675273 were classified under the BR3 subgroup (Figure 4A, Table S3). Combining these five samples, a total of 50,571 cells were obtained, including B cells, CAF, Mono, T cells, EPI_1 (Epithelial with high level of *MMP7* and *ELF3*) and EPI_2 (Epithelial with high level of *HES1*) (Figure 4B, Figure S3). Among these, B cells were significantly enriched in samples derived from BR1 and BR2, while CAFs were significantly enriched in BR3 ones, indicating that the degree of fibroblast infiltration not only affects patient prognosis but also correlates with treatment response (Figure 4C). To elucidate the relationship between CAFs and ovarian cancer treatment response, we further sub-classified CAFs, identifying seven distinct subtypes (Figure 4D), with F_CCL21 and F_RAMP3 significantly enriched in BR1, F_PGF, F_IGFBP4, F_IGFBP7, and F_CXCL12 significantly enriched in BR3, particularly the F_CXCL12 subtype which was almost exclusively found in BR3 (Figure 4E). We then employed gene co-expression analysis to identify key characteristic genes in different CAF subpopulations, resulting in four gene modules, with the blue module showing the highest correlation with the BR3-specific F_CXCL12 subgroup (R=0.88, P<0.001) and the yellow module highly correlated with the BR1-specific F_RAMP3 module (R=0.97, P<0.001) (Figure 4F). Analysis of the hub genes in the blue module revealed that these genes were primarily enriched in pathways related to extracellular matrix remodeling and tube morphogenesis (Figure 4G, S5). Given the significant enrichment of CAFs in BR3, we further explored the cellular communication between these fibroblasts and tumor cells, finding high-intensity interactions between the fibroblast subpopulations F_CXCL12 and F_IGFBP7 and tumor cells in EPI_1 (Figure 4H). Specifically, F_CXCL12 interacts with tumor epithelial cells via ligand-receptor pair CXCL12/CXCR4, and through the secretion of PGF (Figure 4I). These findings suggest that CXCL12-positive CAF cells may be associated with chemotherapy resistant.

**Figure 4.**
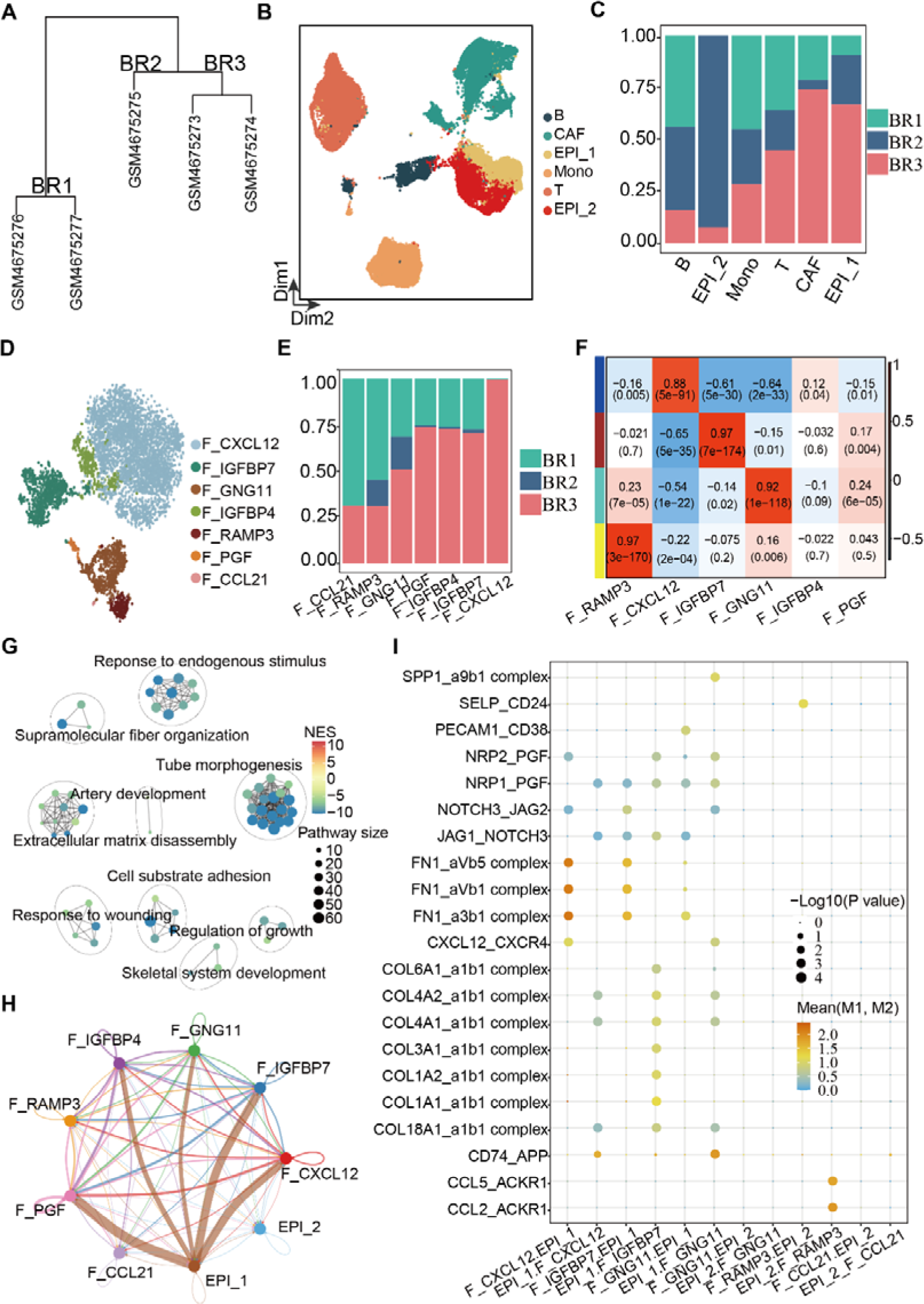
Intra- and inter-heterogeneity of CAF. (A) Tumor phylogenetic tree constructed by hierarchical clustering using the 150 branch genes. (B) UMAP plot showing the major cell types in the dataset GSE154600. (C) Bar plot showing the origins of cell types in three subtypes of branching evolution. (D) UMAP plot showing the subtypes of CAF. (E) Bar plot showing the origins of CAF in the three evolutionary subtypes. (F) WGCNA results showing the gene modules in distinct CAF subtypes. Columns represent cell types. The color from blue to red indicates a low to a high correlation between gene module and cell subtypes (Pearson correlation test). (G) GO enrichment analysis of hub genes of the BR3 enrichment subtype (F_CXCL12). (H) Number of significant ligand-receptor pairs between CAF and epithelial subtypes. The edge width is proportional to the indicated number of ligand-receptor pairs. EPI_1, epithelial subtype with high expression level of MMP7 and ELF3, EPI_3, epithelial subtype with high expression level of HES1 and CD24. (I) Dot plot showing the L–R pairs between CAF and epithelial cells. Rows represent the L–R pairs, and columns represent cell subset–cell subset pairs. The color gradient from black/blue to red indicates mean values of the L–R pairs from low to high, and the circle size indicates the significance of the pairs. p values were calculated via a permutation test using CellChat.

To validate the relationship between fibroblasts and chemotherapy, we further examined the relationship between fibroblasts and treatment response in the other dataset GSE165897 consist of chemotherapy response information (Table S4). Subsequently, using the previously identified 150 characteristic genes, we performed hierarchical evolutionary analysis between samples and conducted permutation test. We found that the platinum-free interval (PFI) values of EOC3, EOC349, EOC540, and EOC87 in the BR3 subgroup were significantly lower than those in the other two groups, indicating significant chemotherapy resistance, while the samples EOC136 and EOC153 in BR1 showed chemotherapy sensitivity, again confirming the high correlation between inter-tumor heterogeneity and treatment response (Figure 5A, Table S4). From this dataset of nine samples, we obtained 17,898 cells, comprising 11 subpopulations, including two fibroblast subpopulations, F_CXCL12 and F_COL1A1, with F_CXCL12 significantly infiltrating in BR3 and F_COL1A1 significantly enriched in BR1 (Figure 5B, 5C, S6). We then re-clustered the fibroblasts, identifying five fibroblast subpopulations, including the F_CXCL12 subgroup (Figure 5D, S6). Using gene co-expression analysis, we also obtained a gene module highly correlated with the F_CXCL12 subgroup (R=0.85, P<0.001, Figure 5E), with characteristic genes primarily enriched in TGF-beta, MAPK, and Cytokine interaction signaling pathways (Figure S7).

**Figure 5.**
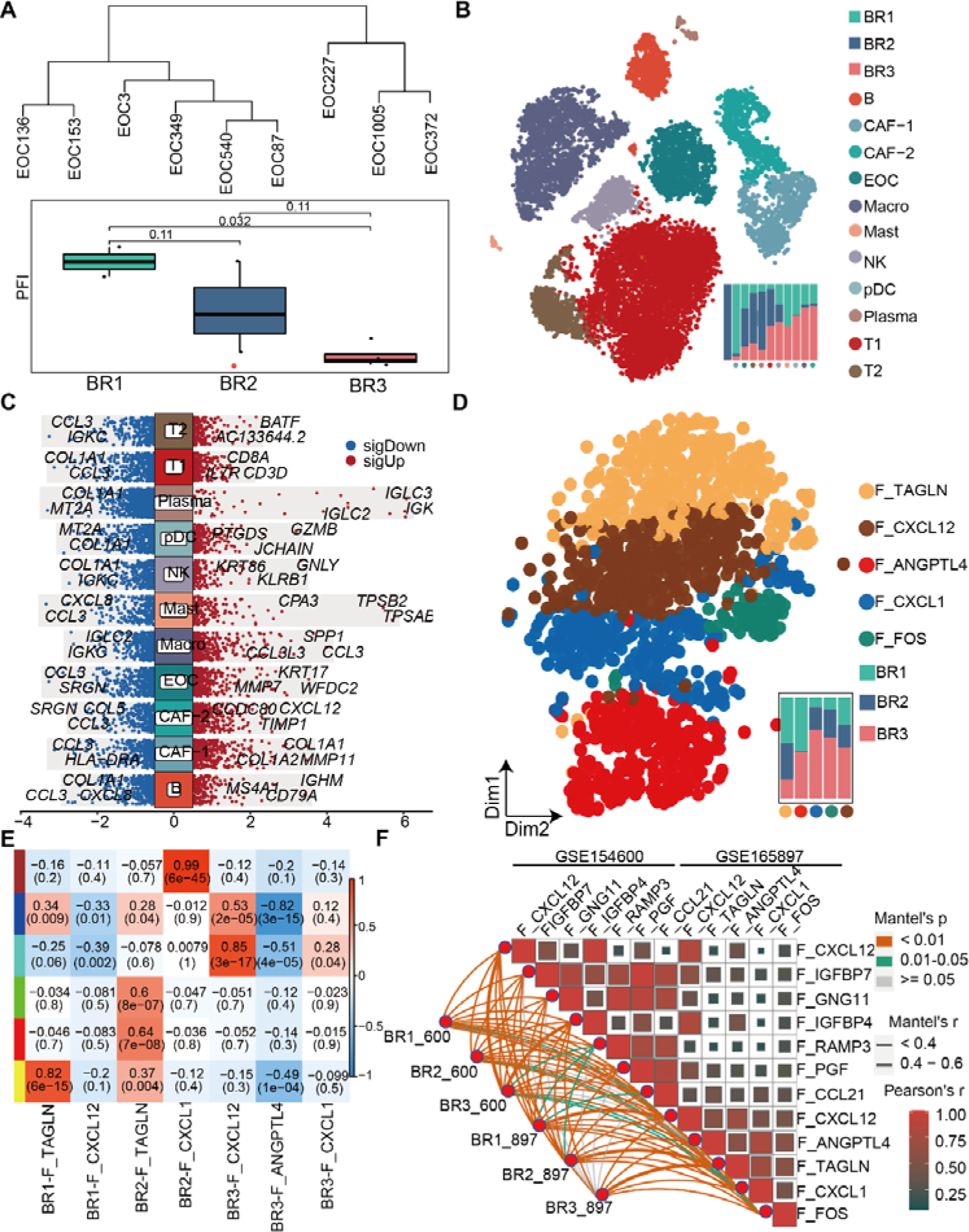
Heterogeneity of CAF is associated with chemotherapy treatment outcomes. (A) Platinum-free interval values in the three tumor evolution branches in dataset GSE165897. (B) UMAP and bar plot showing the major cell types and their origin. (C) Volcano plot showing the differential genes for CAF subtype. Upregulated genes were indicated in red, while downregulated ones in blue. (D) UMAP and bar plot showing the subtype and origin of CAF. (E) WGCNA results showing the gene modules in distinct CAF subtypes in GSE165897. (E) Heatmap showing the CAF subtypes correlation between dataset GSE154600 and GSE165897.

In both datasets, we identified a fibroblast subgroup with high CXCL12 expression, and similarity correlation analysis between subgroups confirmed the high similarity of CXCL12-high fibroblast subgroups in both datasets (Figure 5E).

#### Identification of a gene set affecting prognosis and drug resistance in HGSOC through integrating the signature genes in CXCL12-positive fibroblasts

After identifying a significant correlation between the highly invasive CXCL12+ CAF subpopulation and both treatment responses and prognoses in ovarian cancer. We further conducted a comparative analysis of the WGCNA hub genes results to consolidate the characteristic genes of the CXCL12+ CAF subpopulation, resulting in the identification of 24 shared genes (Table S5). Subsequent Cox regression analysis using the GSE26193 [28] dataset revealed that DCN, CXCL12, and TNFAIP6 were significantly positively associated with poor prognosis in ovarian cancer, with DCN specifically identified as a fibroblast characteristic gene (Figure 6A). Employing these significantly differential genes and Cox regression coefficients, we performed a risk analysis across four additional datasets. The findings indicated that the risk coefficients for the high gene expression groups were notably higher than those for the low expression groups, and both CXCL12 and DCN showed significant positive correlations with poor overall patient prognosis (Figure 6A, 6B). Through Friends analysis, we observed a high correlation of CXCL12 with other genes (Figure 6C). Additionally, using these 24 genes, we applied enrichment analysis in three treatment datasets (GSE33482, GSE114206 [29], and GSE189843 [15]) to further validate the association between CAFs and chemotherapy response (Figure 6D, Table S6). Based on gene expression data from these samples, we categorized them according to their enrichment levels in the 24 genes. The results demonstrated that these 24 genes could effectively stratify samples into high and low groups, with the high CAF subpopulation exhibiting strong drug resistance. Moreover, these 24 genes showed excellent predictive efficacy for drug resistance, with AUC values of 1, 0.89, and 0.83 (Figure 6D). These findings underscore the critical role of fibroblasts in ovarian cancer treatment and prognosis, and through further feature extraction, we have identified a reduced set of genes that are indicative of treatment response.

**Figure 6.**
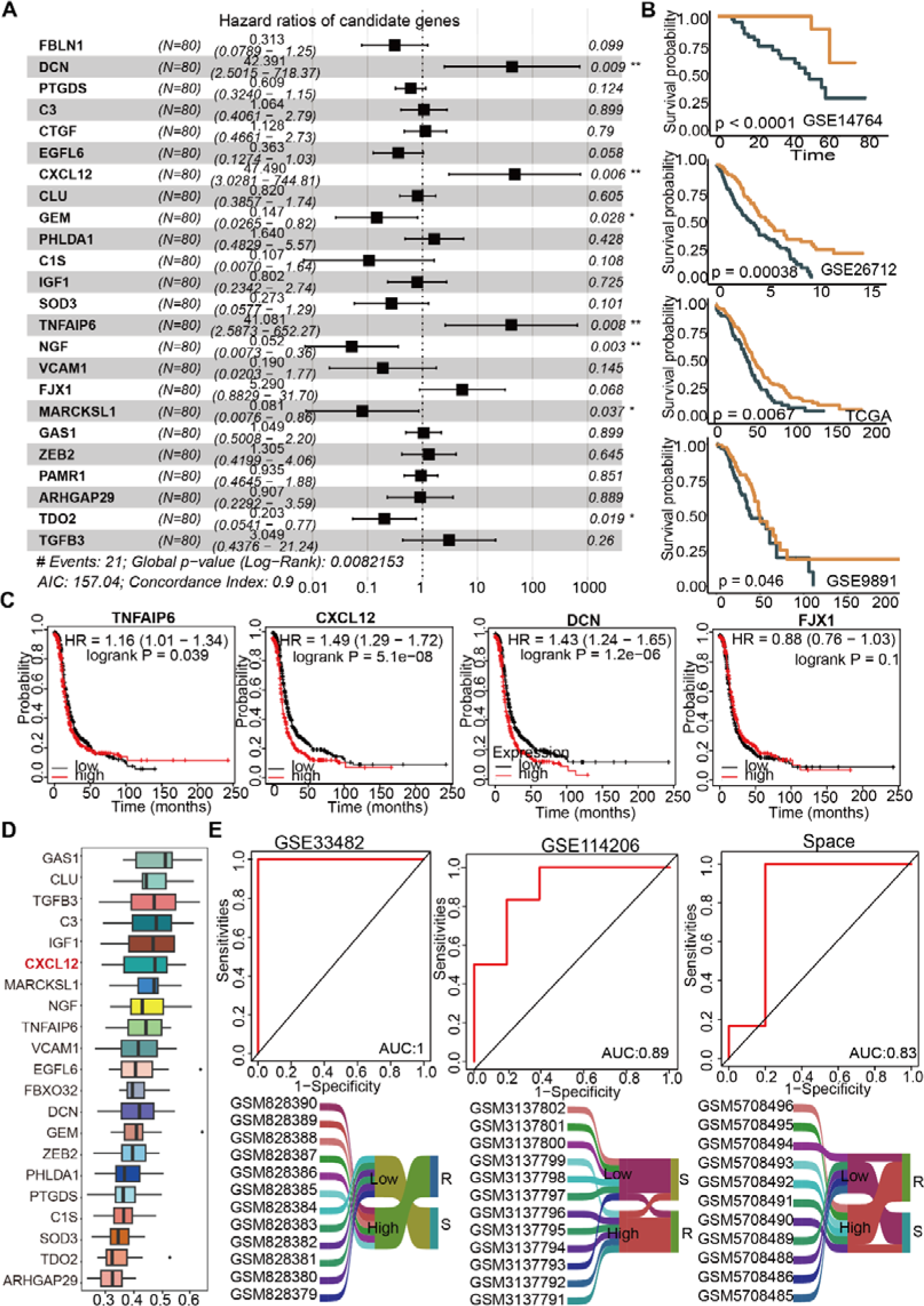
24-genes of CXCL12-positive fibroblasts were correlated with prognosis and drug resistance in HGSOC. (A). Forest plot showing the risk prognosis results from 24 shared CXCL12-positive fibroblasts obtained from two single-cell samples via COX regression. (B) Overall survival curves showing the prognosis results with different level of CAF risk score in the four HGSOC cohorts using the 24 genes. Statistical significance was calculated using the log-rank test. (C) Overall survival curves showing the prognosis result of the high HR scores genes. (D) Box plot showing the friends analysis results. (E) AUC and Sankey diagram show the prediction of chemotherapy resistance using 24 CXCL12-CAF genes.

#### The spatial distribution characteristics of CXCL12-positive fibroblasts affect the clinical prognostic results of HGSOC

Given the strong correlation between CXCL12-positive fibroblasts and the treatment outcomes and prognoses of high-grade serous ovarian cancer, as well as their interaction with tumor cells via CXCR4, we further elucidated the interaction mechanisms of CXCL12-positive fibroblasts in vitro and in spatial transcriptomic data. In our in vitro experiments, we silenced the CXCL12 receptor gene CXCR4 in the ovarian cancer cell line SKOV3 (Figure 7A). As shown by qPCR, all three siRNA knockdown fragments significantly suppressed CXCR4 mRNA expression levels in SKOV3 cells (p < 0.05, Figure 7A). Subsequent addition of exogenous CXCL12 protein to both the control group and the si-R2 group demonstrated that CXCR4 knockdown significantly inhibited cell viability compared to the si-NC control group (p < 0.05, Figure 7B, 7C). Considering that fibroblasts and tumor cells interact through cellular communication, and the intensity of this communication is highly related to the spatial positioning of genes, we further analyzed the spatial distribution characteristics of fibroblasts and tumor cells within a treatment response-associated spatial transcriptomic dataset. Using the inferCNV method, we identified tumor regions and the surrounding cellular environments. Results from a chemotherapy-resistant sample (GSM6506110) showed high CXCR4 expression in the tumor region (neb_4), with a significant enrichment of fibroblasts in the surrounding area (cluster4) that highly expressed CXCL12 and DCN (Figure 7C, Figure S8). In contrast, in a chemotherapy-sensitive sample (Table S7, Figure S9), although fibroblasts were also enriched, they did not form a spatial neighborhood gene interaction pair of CXCL12-CXCR4 (Figure 7D). Finally, in our treatment response-associated clinical samples, we also validated the local spatial adjacency of CXCL12/DCN/KRT19 in chemotherapy-resistant samples (Figure 7E).

**Figure 7.**
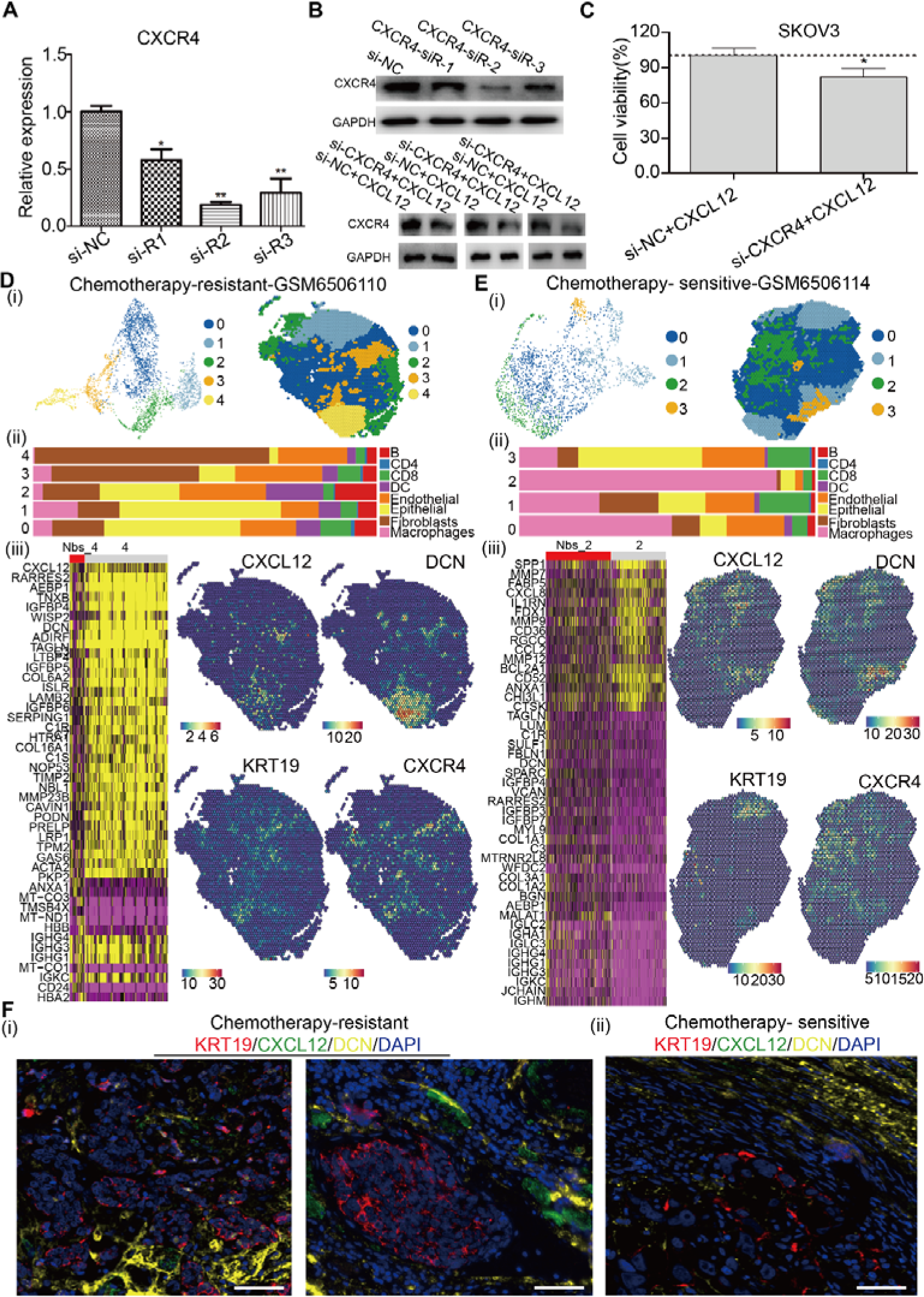
Function and spatial distribution characteristics of CXCL12-positive fibroblasts. (A) The bar chart shows the silencing effect of the CXCL12 receptor gene CXCR4(ns, no significance, *P < 0.05, **P < 0.01, ***P < 0.001, t test). (B) The WB results show the silencing effect of CXCR4, as well as the expression levels of CXCR4 protein in the control group and the CXCR4 silenced group after the addition of exogenous CXCL12 protein. (C) CCK-8 result showing that silencing CXCR4 significantly inhibits the viability of tumor cells (*, P < 0.05, t.test). Clustering and spatial distribution (i), cell type composition in each cluster (ii), and gene profile around the tumor boundary (iii) in chemotherapy-resistant samples (D) and chemotherapy-sensitive samples (E). (F) The multiplex immunofluorescence results show the spatial proximity relationship between fibroblasts and tumor cells in chemotherapy-sensitive and chemotherapy-resistant samples. The scale bars represent 50 µm

## Discussion

Solid-organ malignancies exhibit heterogeneity that is evolutionarily favored to enhance tumor cell survival and drug resistance, likely via a set of molecular mechanisms [30]. Especially the intratumor heterogeneity evolves spatially and temporally in the tumor development and reprogrammed the features of TME in the progression [31]. Defining tumor clonality and its evolutionary path is essential to understand the mechanics of tumor cells’ collective behavior and potential driving factors [32, 33]. Tumors harbor a multitude of genetic and epigenetic alterations, many of which do not directly correlate with observable phenotypic changes, complicating the interpretation of their functional impact on tumor evolution [34]. Single-cell and spatial transcriptomic offers a powerful tool to delineate the distinct functional roles of various cell types within tumors, revealing their unique cellular states and phenotypic characteristics across different organs [35–37]. By focusing on the transcriptional programs of tumor subclones, researchers can better capture the functional diversity that arises from genetic heterogeneity, providing insights into how these subclones adapt and contribute to tumor functional heterogeneity. Tracing tumor subclones via their transcriptional programs, as a means to closely reflect their phenotypic characteristics, emerges as a vital strategy to understand the functional diversity stemming from genetic heterogeneity and environmental adaptation [38, 39]. In this study, we integrated single-cell, bulk and spatial transcriptomic data, and elucidated the intratumoral heterogeneity including tumor and TMEs cells of HGSC, explore their evolutionary patterns intra- and inter heterogeneity, and identify critical factors influencing tumor prognosis and therapeutic responses.

Single-cell data processing faces challenges with large-scale datasets, necessitating efficient computational and statistical methods to handle data sparsity and ensure informative analysis [40]. Batch effects between datasets require robust correction techniques to guarantee accurate cross-dataset comparisons and integrations [40]. Determining the optimal number of clusters is essential for the analysis of intratumor heterogeneity, yet this process is complicated by the lack of clear standards [41]. In this study, we initially identified intratumor heterogeneity using the results from Seurat’s initial clustering result combined with CNV data. We extracted tumor-associated feature genes and identified “representative cells” that highly reflect the characteristics of different tumor clones from subpopulations using the “TF-IDF” method of text mining. We then further analyzed the intratumoral heterogeneity of these cells. This approach ensures the extraction of the most accurate and essential tumor clone information while minimizing computational resource consumption. Additionally, it mitigates the impact of high sparsity in single-cell transcriptome data on data analysis. Furthermore, we utilized the previously identified “representative cells”, and determined the number automatically and therefor confirm the intratumor heterogeneity using SC3 consensus clustering and SVM methods. This strategy addresses the subjectivity associated with the manual definition of cluster numbers. Finally, by obtaining precise intratumoral heterogeneity and related feature genes, we constructed a tumor evolution model based on their expression levels and analyzed the characteristics of intratumoral and intertumoral heterogeneity in HGSOC. Through the integrated analysis of intratumoral and intertumoral heterogeneity, we can more comprehensively elucidate the roles of tumor and microenvironmental cells in the prognosis prediction and chemoresistance of HGSOC. This provides a novel strategy for interpreting correlation between intra- and inter-tumor heterogeneity using expression data.

Employing the aforementioned strategy, we analyzed and validated across multiple datasets that HGSOC patients can be classified into three evolutionary branches: BR1, BR2, and BR3. Notable heterogeneity was observed within each branch concerning the quantity and composition of tumor and microenvironment cells. The BR1 branch was predominated by tumor cells and resembled cold tumors, BR2 was infiltrated with a higher number of B and T cells resembling hot tumors, and BR3 was heavily infiltrated by fibroblasts, akin to immunologically excluded tumors [27]. By correlating tumor evolution with prognosis and resistance to therapy, we found that a high infiltration of fibroblasts was associated with a tumor cell subpopulation enriched in cell cycle pathways, low CD8 cell activity, increased cell proliferation, and malignant transformation in cell morphology, resulting in significantly worse prognosis and therapy resistance. Fibroblasts can be broadly classified into myofibroblasts, TGF-β-driven cancer-associated fibroblasts, inflammatory fibroblasts, and antigen-presenting fibroblasts, and are highly associated with poor prognosis in various cancers [20, 42, 43]. In HGSOC, fibroblasts can form a dense physical barrier around tumor cells, impede the infiltration of immune cells, and promote tumor cell proliferation through the secretion of cytokines [44, 45]. Combing with WGCNA, we further identified a clinical predictive model comprising 24 genes and discovered that fibroblasts with high CXCL12 expression are significantly related to poor tumor prognosis and chemotherapy resistance. The differences in the spatial distribution of cells, particularly the characteristics and functions of cells near the tumor boundary, play a crucial role in the development of tumor heterogeneity [25]. To this end, we developed a bioinformatics analytical workflow for tumor boundary identification and found that this type of fibroblast significantly infiltrates the margins of chemotherapy-resistant tumors. Spatial interactions between cell clusters may have a more profound impact on chemo responsiveness than the composition of the clusters alone. In HGSOC, fibroblasts can interact with other cell types through various pathways, ultimately contributing to chemotherapy resistance [43]. CXCL12+ CAFs could promote cancer cell migration and invasion and upregulate PDL1 in bladder and pancreatic cancer [46, 47]. They can also attract monocytes through the CXCL12/CXCR4 pathway and induce their differentiation into M2 macrophages, which leads to enhanced tumor cell proliferation and reduced apoptosis in oral squamous cell carcinoma [48]. In this study, by analyzing the evolutionary pathways of tumor and microenvironment cells, we determined that CXCL12-expressing fibroblasts, through spatial proximity interactions with tumor cells, influence patient prognosis and therapeutic outcomes, offering a new perspective for the prevention and treatment of HGSOC.

## Acknowledgements

The authors thank Dr. Mengyun Ke for the sample collection and storage.

## Availability of data

The datasets analyzed in this study are available from the gene expression omnibus (GEO) repository under the accession numbers in supplementary tables.

## Author contributions

The study design and supervision were conducted by Yongjun Guo, Jun Yang, Jun Li, and Tingjie Wang. Data analysis, in vitro experiments, and manuscript writing were performed by Tingjie Wang, Lingxi Tian, and Ruitao Long. Clinical data collection, sorting, analysis, manuscript proofreading, and multicolor immunofluorescence staining analysis were carried out by Bing Wei, Cuiyun Zhang, Bo Wang, Yougai Zhang. Yougai Zhang and Xiaofei Zhu verified the patients’ clinical data and evaluated the therapeutic responses. The manuscript has been reviewed and approved by all authors.

## Reference

1. Coburn SB, Bray F, Sherman ME, Trabert B. International patterns and trends in ovarian cancer incidence, overall and by histologic subtype. Int J Cancer. 2017; 140: 2451–60.

2. Chowdhury S, Kennedy JJ, Ivey RG, Murillo OD, Hosseini N, Song X, et al. Proteogenomic analysis of chemo-refractory high-grade serous ovarian cancer. Cell. 2023; 186: 3476–98 e35.

3. Zheng X, Wang X, Cheng X, Liu Z, Yin Y, Li X, et al. Single-cell analyses implicate ascites in remodeling the ecosystems of primary and metastatic tumors in ovarian cancer. Nat Cancer. 2023; 4: 1138–56.

4. Matthews BG, Bowden NA, Wong-Brown MW. Epigenetic Mechanisms and Therapeutic Targets in Chemoresistant High-Grade Serous Ovarian Cancer. Cancers (Basel). 2021; 13.

5. Silva R, Glennon K, Metoudi M, Moran B, Salta S, Slattery K, et al. Unveiling the epigenomic mechanisms of acquired platinum-resistance in high-grade serous ovarian cancer. Int J Cancer. 2023; 153: 120–32.

6. Liu J, Dang H, Wang XW. The significance of intertumor and intratumor heterogeneity in liver cancer. Exp Mol Med. 2018; 50: e416.

7. Olbrecht S, Busschaert P, Qian J, Vanderstichele A, Loverix L, Van Gorp T, et al. High-grade serous tubo-ovarian cancer refined with single-cell RNA sequencing: specific cell subtypes influence survival and determine molecular subtype classification. Genome Med. 2021; 13: 111.

8. Zhu JW, Charkhchi P, Akbari MR. Potential clinical utility of liquid biopsies in ovarian cancer. Mol Cancer. 2022; 21: 114.

9. Zellmer V R ZS. Evolving concepts of tumor heterogeneity. Cell & bioscience. 2014; 4: 1–8.

10. Polyak K. Tumor heterogeneity confounds and illuminates: a case for Darwinian tumor evolution. Nat Med. 2014; 20: 344–6.

11. Cordani M, Dando I, Ambrosini G, Gonzalez-Menendez P. Signaling, cancer cell plasticity, and intratumor heterogeneity. Cell Commun Signal. 2024; 22: 255.

12. Ciriello G, Magnani L, Aitken SJ, Akkari L, Behjati S, Hanahan D, et al. Cancer Evolution: A Multifaceted Affair. Cancer Discov. 2024; 14: 36–48.

13. de Visser KE, Joyce JA. The evolving tumor microenvironment: From cancer initiation to metastatic outgrowth. Cancer Cell. 2023; 41: 374–403.

14. Ferri-Borgogno S, Zhu Y, Sheng J, Burks JK, Gomez JA, Wong KK, et al. Spatial Transcriptomics Depict Ligand-Receptor Cross-talk Heterogeneity at the Tumor-Stroma Interface in Long-Term Ovarian Cancer Survivors. Cancer Res. 2023; 83: 1503–16.

15. Stur E, Corvigno S, Xu M, Chen K, Tan Y, Lee S, et al. Spatially resolved transcriptomics of high-grade serous ovarian carcinoma. iScience. 2022; 25: 103923.

16. Khatib S, Pomyen Y, Dang H, Wang XW. Understanding the Cause and Consequence of Tumor Heterogeneity. Trends Cancer. 2020; 6: 267–71.

17. Iacobuzio-Donahue CA, Litchfield K, Swanton C. Intratumor heterogeneity reflects clinical disease course. Nature Cancer. 2020; 1: 3–6.

18. Stuart T, Butler A, Hoffman P, Hafemeister C, Papalexi E, Mauck WM, 3rd, et al. Comprehensive Integration of Single-Cell Data. Cell. 2019; 177: 1888–902 e21.

19. Lun AT, McCarthy DJ, Marioni JC. A step-by-step workflow for low-level analysis of single-cell RNA-seq data with Bioconductor. F1000Res. 2016; 5: 2122.

20. Wang T, Xu C, Zhang Z, Wu H, Li X, Zhang Y, et al. Cellular heterogeneity and transcriptomic profiles during intrahepatic cholangiocarcinoma initiation and progression. Hepatology. 2022; 76: 1302–17.

21. Kiselev VY, Kirschner K, Schaub MT, Andrews T, Yiu A, Chandra T, et al. SC3: consensus clustering of single-cell RNA-seq data. Nat Methods. 2017; 14: 483–6.

22. Tosches M A YTM, Naumann R K. Evolution of pallium, hippocampus, and cortical cell types revealed by single-cell transcriptomics in reptiles. Science. 2018; 360: 881–8.

23. Zhang K, Erkan EP, Jamalzadeh S, Dai J, Andersson N, Kaipio K, et al. Longitudinal single-cell RNA-seq analysis reveals stress-promoted chemoresistance in metastatic ovarian cancer. Sci Adv. 2022; 8: eabm1831.

24. Yu G, Li F, Qin Y, Bo X, Wu Y, Wang S. GOSemSim: an R package for measuring semantic similarity among GO terms and gene products. Bioinformatics. 2010; 26: 976–8.

25. Xun Z, Ding X, Zhang Y, Zhang B, Lai S, Zou D, et al. Reconstruction of the tumor spatial microenvironment along the malignant-boundary-nonmalignant axis. Nat Commun. 2023; 14: 933.

26. Aran D, Hu Z, Butte AJ. xCell: digitally portraying the tissue cellular heterogeneity landscape. Genome Biol. 2017; 18: 220.

27. Hegde PS, Chen DS. Top 10 Challenges in Cancer Immunotherapy. Immunity. 2020; 52: 17–35.

28. Gentric G, Kieffer Y, Mieulet V, Goundiam O, Bonneau C, Nemati F, et al. PML-Regulated Mitochondrial Metabolism Enhances Chemosensitivity in Human Ovarian Cancers. Cell Metabolism. 2019; 29: 156–73.e10.

29. Veskimae K, Scaravilli M, Niininen W, Karvonen H, Jaatinen S, Nykter M, et al. Expression Analysis of Platinum Sensitive and Resistant Epithelial Ovarian Cancer Patient Samples Reveals New Candidates for Targeted Therapies. Transl Oncol. 2018; 11: 1160–70.

30. Ramon YCS, Sese M, Capdevila C, Aasen T, De Mattos-Arruda L, Diaz-Cano SJ, et al. Clinical implications of intratumor heterogeneity: challenges and opportunities. J Mol Med (Berl). 2020; 98: 161–77.

31. Marusyk A, Janiszewska M, Polyak K. Intratumor Heterogeneity: The Rosetta Stone of Therapy Resistance. Cancer Cell. 2020; 37: 471–84.

32. Alizadeh AA, Aranda V, Bardelli A, Blanpain C, Bock C, Borowski C, et al. Toward understanding and exploiting tumor heterogeneity. Nat Med. 2015; 21: 846–53.

33. Aizarani N, Saviano A, Sagar, Mailly L, Durand S, Herman JS, et al. A human liver cell atlas reveals heterogeneity and epithelial progenitors. Nature. 2019; 572: 199–204.

34. Ma L, Wang L, Khatib SA, Chang CW, Heinrich S, Dominguez DA, et al. Single-cell atlas of tumor cell evolution in response to therapy in hepatocellular carcinoma and intrahepatic cholangiocarcinoma. J Hepatol. 2021; 75: 1397–408.

35. Tasic B, Yao Z, Graybuck LT, Smith KA, Nguyen TN, Bertagnolli D, et al. Shared and distinct transcriptomic cell types across neocortical areas. Nature. 2018; 563: 72–8.

36. Gonzalez-Silva L, Quevedo L, Varela I. Tumor Functional Heterogeneity Unraveled by scRNA-seq Technologies. Trends Cancer. 2020; 6: 13–9.

37. Ru B, Huang J, Zhang Y, Aldape K, Jiang P. Estimation of cell lineages in tumors from spatial transcriptomics data. Nat Commun. 2023; 14: 568.

38. Solary E, Laplane L. The role of host environment in cancer evolution. Evolutionary Applications. 2020; 13: 1756–70.

39. S A, Chakraborty A, Patnaik S. Clonal evolution and expansion associated with therapy resistance and relapse of colorectal cancer. Mutat Res Rev Mutat Res. 2022; 790: 108445.

40. Lahnemann D, Koster J, Szczurek E, McCarthy DJ, Hicks SC, Robinson MD, et al. Eleven grand challenges in single-cell data science. Genome Biol. 2020; 21: 31.

41. Yu L, Cao Y, Yang JYH, Yang P. Benchmarking clustering algorithms on estimating the number of cell types from single-cell RNA-sequencing data. Genome Biol. 2022; 23: 49.

42. Zhang L, Cascio S, Mellors JW, Buckanovich RJ, Osmanbeyoglu HU. Single-cell analysis reveals the stromal dynamics and tumor-specific characteristics in the microenvironment of ovarian cancer. Commun Biol. 2024; 7: 20.

43. Hussain A, Voisin V, Poon S, Karamboulas C, Bui NHB, Meens J, et al. Distinct fibroblast functional states drive clinical outcomes in ovarian cancer and are regulated by TCF21. J Exp Med. 2020; 217.

44. Mao X, Xu J, Wang W, Liang C, Hua J, Liu J, et al. Crosstalk between cancer-associated fibroblasts and immune cells in the tumor microenvironment: new findings and future perspectives. Mol Cancer. 2021; 20: 131.

45. Cai J, Tang H, Xu L, Wang X, Yang C, Ruan S, et al. Fibroblasts in omentum activated by tumor cells promote ovarian cancer growth, adhesion and invasiveness. Carcinogenesis. 2012; 33: 20–9.

46. Feig C, Jones JO, Kraman M, Wells RJ, Deonarine A, Chan DS, et al. Targeting CXCL12 from FAP-expressing carcinoma-associated fibroblasts synergizes with anti-PD-L1 immunotherapy in pancreatic cancer. Proc Natl Acad Sci U S A. 2013; 110: 20212–7.

47. Zhang Z, Yu Y, Zhang Z, Li D, Liang Z, Wang L, et al. Cancer-associated fibroblasts-derived CXCL12 enhances immune escape of bladder cancer through inhibiting P62-mediated autophagic degradation of PDL1. J Exp Clin Cancer Res. 2023; 42: 316.

48. Li X, Bu W, Meng L, Liu X, Wang S, Jiang L, et al. CXCL12/CXCR4 pathway orchestrates CSC-like properties by CAF recruited tumor associated macrophage in OSCC. Exp Cell Res. 2019; 378: 131–8.

